# The impact of COVID-19 non-pharmaceutical interventions on the lived experiences of people living in Thailand, Malaysia, Italy and the United Kingdom: a cross-country qualitative study

**DOI:** 10.1101/2021.05.13.21257162

**Authors:** Mira L. Schneiders, Bhensri Naemiratch, Phaik Kin Cheah, Giulia Cuman, Tassawan Poomchaichote, Supanat Ruangkajorn, Silvia Stoppa, Anne Osterrieder, Phee-Kheng Cheah, Darlene Ongkili, Wirichada Pan-ngum, Constance R.S. Mackworth-Young, Phaik Yeong Cheah

## Abstract

This qualitative study explores the impact of non-pharmaceutical interventions (NPIs) on lived experiences during the first wave of the COVID-19 pandemic on people’s lives in Thailand, Malaysia, Italy and the United Kingdom. A total of 86 interviews were conducted with members of the public, including healthcare workers. Participants across countries held strong views on NPIs, with many feeling measures lacked clarity. Most participants reported primarily negative impacts of NPIs on their lives, including through separation, isolation and grief over missed milestones; work-related challenges and income loss; and poor mental health and wellbeing. Nonetheless, many also experienced inadvertent positive consequences, including more time at home to focus on what they most valued in life; a greater sense of connectedness; and benefits to working life. Commonly employed coping strategies focused on financial coping; psycho-emotional coping; social coping and connectedness; reducing and mitigating risks; and limiting exposure to the news. Importantly, the extent to which participants’ lived experiences were positive or negative, and their ability to cope was underpinned by individual, social and economic factors. In order to mitigate negative and unequal impacts of NPIs, COVID-19 policies will benefit from paying closer attention to the social, cultural and psychological—not just biological—vulnerabilities to, and consequences of public health measures.

## Introduction

> *“We are like islands in the sea, separate on the surface but connected in the deep*.*”* William James

In 2020, countries around the world implemented a range of containment measures using non-pharmaceutical interventions (NPIs) to prevent the transmission of SARS-CoV-2 (COVID-19), which to date has resulted in 113,467,303 confirmed cases, including 2,520,550 deaths worldwide [1]. These included social distancing measures, such as contact tracing, quarantine, closure of schools, work places and non-essential shops; travel-related measures such as travel restrictions, screening and border closure; and personal protective measures such as face masks, hand hygiene and respiratory etiquette [2]. Evidence supports the effectiveness of these measures in reducing COVID-19 transmission, mortality and healthcare demand [3-6].

Despite this evidence, ethical considerations need to account for the economic, social and psychological impacts of NPIs on people’s lives [7]. For this, it is vital to understand how NPIs are experienced by the public. A systematic review and synthesis of qualitative studies on public perceptions on NPIs in the context of respiratory infections found that perceptions were impacted by assessments about their necessity, efficacy, acceptability, and feasibility [8]. Furthermore, the potential to attract stigma, perceptions about the emotional, personal and societal economic costs, and cultural unacceptability were identified as disadvantages of social distancing, isolation and quarantine [8].

The importance of focusing on mental health amidst this pandemic has been underscored by the emergence of evidence highlighting the negative psychological impacts of NPIs [9]. A recent review on the psychological impact of quarantine identified post-traumatic stress symptoms, confusion, and anger as negative effects, and found that longer quarantine duration, infection fears, frustration, boredom, inadequate information and supplies, financial loss, and stigma were important stressors [10]. While these findings do not suggest that NPIs, like quarantine and isolation should not be employed, the adverse psychological impacts they may cause need to be weighed against likely benefits [10].

Emerging evidence also suggests that the use of NPIs in the context of COVID-19 has had unequal effects on society, including across and within countries [11, 12]. Vulnerable populations in particular, including migrants, people working in the informal economy, homeless people, older people and people living with disabilities, may be disproportionately affected by the adverse impacts of NPIs [7, 13]. Vulnerable individuals may be less able to comply with NPIs, due to underlying inequalities in the distribution of resources, wealth and power [13-15]. For example, they may be less able to practice hand hygiene (if lacking access to basic sanitation), wear masks (due to access and affordability issues) or adopt recommended social distancing (when living in crowded conditions, relying on public transport, working in high public contact jobs or being unable to access sick leave) [13-15]. This, in turn is likely to exacerbate existing health, economic and social inequalities.

Understanding the social, economic and mental health impacts of prolonged exposure to social distancing and isolation during the COVID-19 pandemic has been identified as a priority for multidisciplinary COVID-19 research [16]. The importance of doing this research “together with people with lived experience” has also been highlighted [17]. Qualitative research in particular can help to gain a rich and comprehensive understanding of lived experience [18]. Knowledge generated using social science methods is thus crucial for generating insights that can inform the development of more effective and equitable public health interventions in light of infectious disease threats [19].

To date, little qualitative research has been conducted to understand public perceptions and lived experiences of NPIs. A focus group study conducted in the UK found that feelings of anxiety, depression and loss were common in response to COVID-19 social distancing and isolation [20]. However, the study did not include healthcare workers (HCW) or people at higher-risk of severe illness from COVID-19, such as those with underlying chronic conditions or over the age of 60 years [21], and so little is known about how these groups are impacted by NPIs. While no qualitative studies have directly studied the impact of NPIs on HCW, existing quantitative evidence suggests that HCW are at greater risk of experiencing psychological distress during the COVID-19 outbreak, including symptoms of depression, anxiety and PTSD [22-25]. Furthermore, to our knowledge, no qualitative study has been conducted comparing lived experiences of NPIs across different countries and world regions. Cross-national studies are useful because they can facilitate understanding of common issues [26] and generate deeper insights into the phenomena being studied [27]. This study therefore aims to help address these gaps. Using in-depth interviews, we seek to explore and compare the lived experiences, coping strategies and views of COVID-19 NPIs among the public and HCW across four countries, namely Thailand, Malaysia, Italy and the United Kingdom (UK).

## Materials and methods

### Study design

In-depth, semi-structured interviews were conducted with 86 individuals residing in four countries: Thailand, Malaysia, Italy and the UK between May 2 and August 8 2020 (TH: 8.05-21.07.2020; MY: 2.05-4.07.2020; IT: 13.05-04.08.2020; UK: 14.05-23.07.2020). Countries were selected based on the implementation of COVID-19 public health measures (NPIs) during the study period and the existence of prior research collaborations, which enabled and facilitated timely implementation of the study. Here we describe results from a qualitative study, which was part of a mixed-methods study (SEB-COV) that also included an online survey [28]. The quantitative results are reported elsewhere [11].

### Theoretical framework

This study used a phenomenological approach, focused on “the study of an individual’s lived experiences within the world” [29]. In-depth interviews were used to encourage participants to narrate their own lived experiences of government imposed NPIs. Thematic analysis was used to code data into themes and sub-themes, based on the Framework Method [30].

### COVID-19 government response in the four countries

Over the study period, Thailand, Malaysia, Italy and the UK were under varying degrees of lockdown to prevent the transmission of COVID-19, which included closures of workplaces and schools, restrictions on movement and travel, and social distancing guidelines. The Oxford COVID-19 Government Response Tracker (OxCGRT) [31] tracks government responses to COVID-19 from more than 180 countries on standardized indicators. Data are aggregated to produce a daily ‘Stringency Index’ (SI), on a scale from 0-100, with 100 indicating the strictest public health response. Fig 1 visualises the SI of the four governments over the study period. For example, Italy experienced the strictest restrictions at the beginning of May (SI=93), followed by easing of restrictions in early June (SI=44) and subsequent tightening. In Thailand, the SI was high at the start of the study (SI=77) and decreased at the end of June (SI=53). The pattern was similar in Malaysia. Restrictions in the UK remained high throughout (SI=69-76).

**Fig 1.**
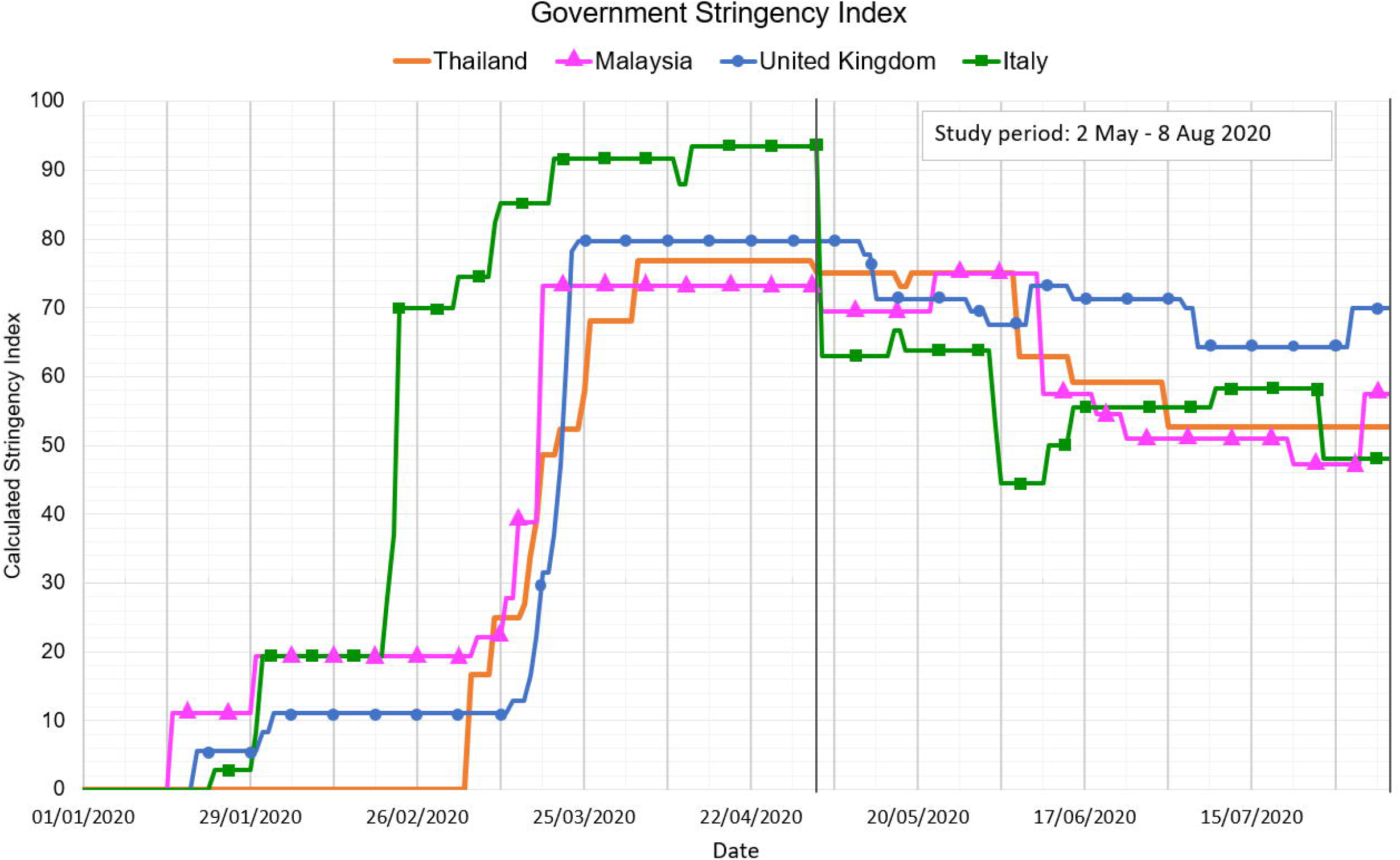
Government Stringency Index. SI during the first wave of COVID-19 NPIs for Thailand, Malaysia, Italy and the UK.

### Participant selection

All participants talking part in the SEB-COV online survey [28] were invited to take part in the qualitative arm of the study. Additionally, in each country, participants were recruited through existing research and organisational networks, social media channels (Facebook, Twitter), and targeted social media advertising (Facebook adverts). To encourage greater diversity among the sample, key community organisations and institutions working with vulnerable, hard-to-reach, or high-risk populations (e.g., entertainment workers, family caregivers, frontline workers, older people) were contacted via email and asked to disseminate information about the study through their networks.

Individuals were provided with information about the study and asked to register their interest using a short socio-demographic recruitment survey online, administered using the ‘JISC online surveys’ platform [32]. Across countries, a total of 276 participants registered their interest via the recruitment survey (TH: 34; MY: 52; IT: 34; UK: 156). Among these, participants were selected by each country team using purposive sampling to gain a maximum variation sample, based on six characteristics (i.e., gender, age, number of household members, self-perceived level of risk, level of education and occupation). Selected individuals were invited to take part in an interview via email or telephone (total 127, TH: 29; MY: 24; IT: 34; UK: 40). Of those contacted, 91 individuals agreed to participate, with five dropping out prior to the interview. A total of 86 interviews were therefore conducted across the four countries.

Participants in Thailand received ฿200 THB (USD 7), and in the UK were provided a £10 electronic supermarket voucher to compensate for their time. No compensation was provided in Italy and Malaysia, in accordance with local research practices.

### Participant characteristics

Overall, 51% of all participants were female, 43% male and 6% identified as other genders (Table 1). Participant age ranged from 18-84 years (IT:20-65; MY 19-84; TH 18-74; UK:24-80 years), with a total of 12.8% (n=11) aged 65 years or older. Participants varied in their levels of education (5% primary, 31% secondary, 64% tertiary), household size (range: 1-12 people) and occupations (15% HCW, 85% other). Across all countries, 41% of participants perceived themselves at low, 37% at medium and 22% at high risk of COVID-19.

**Table 1.**
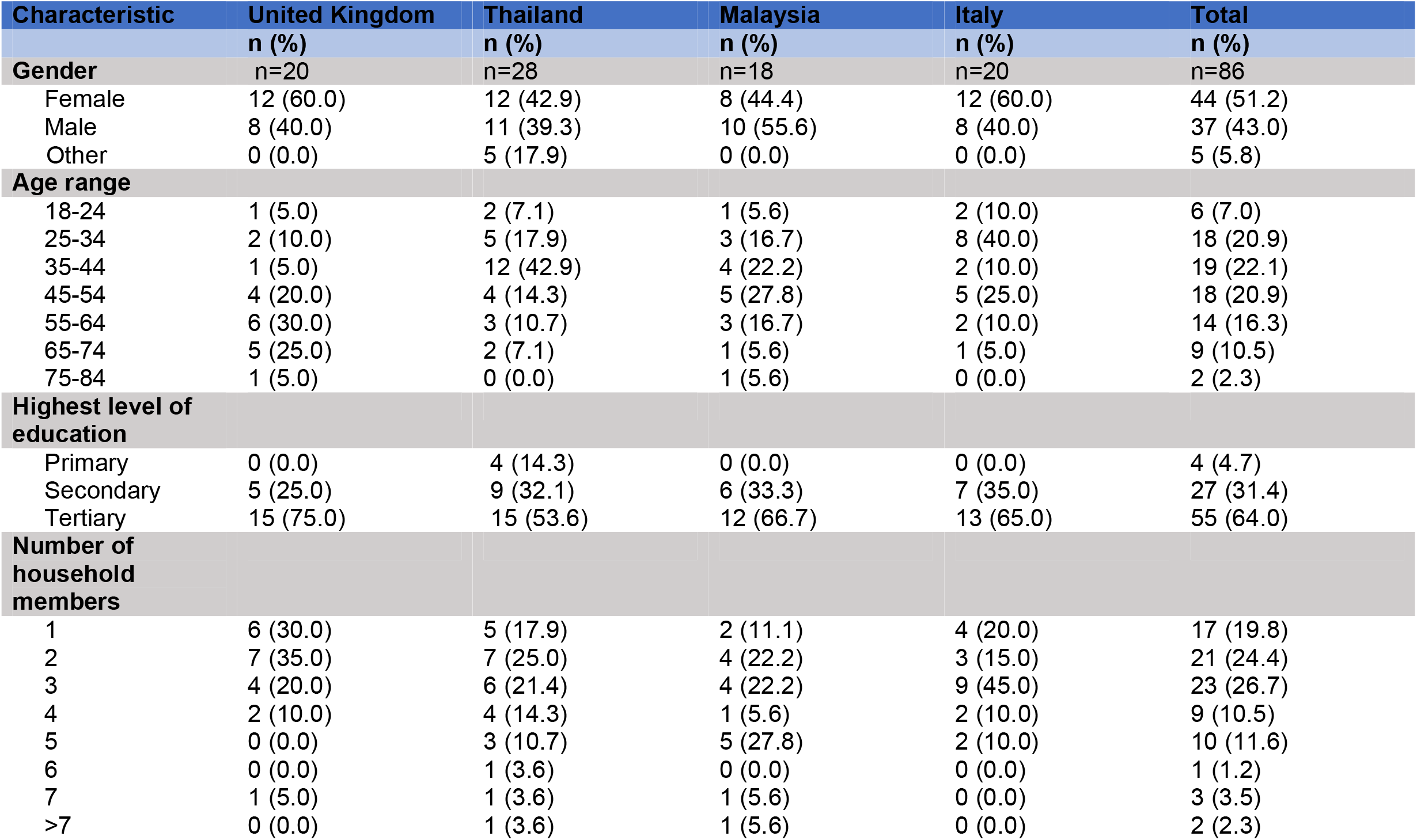
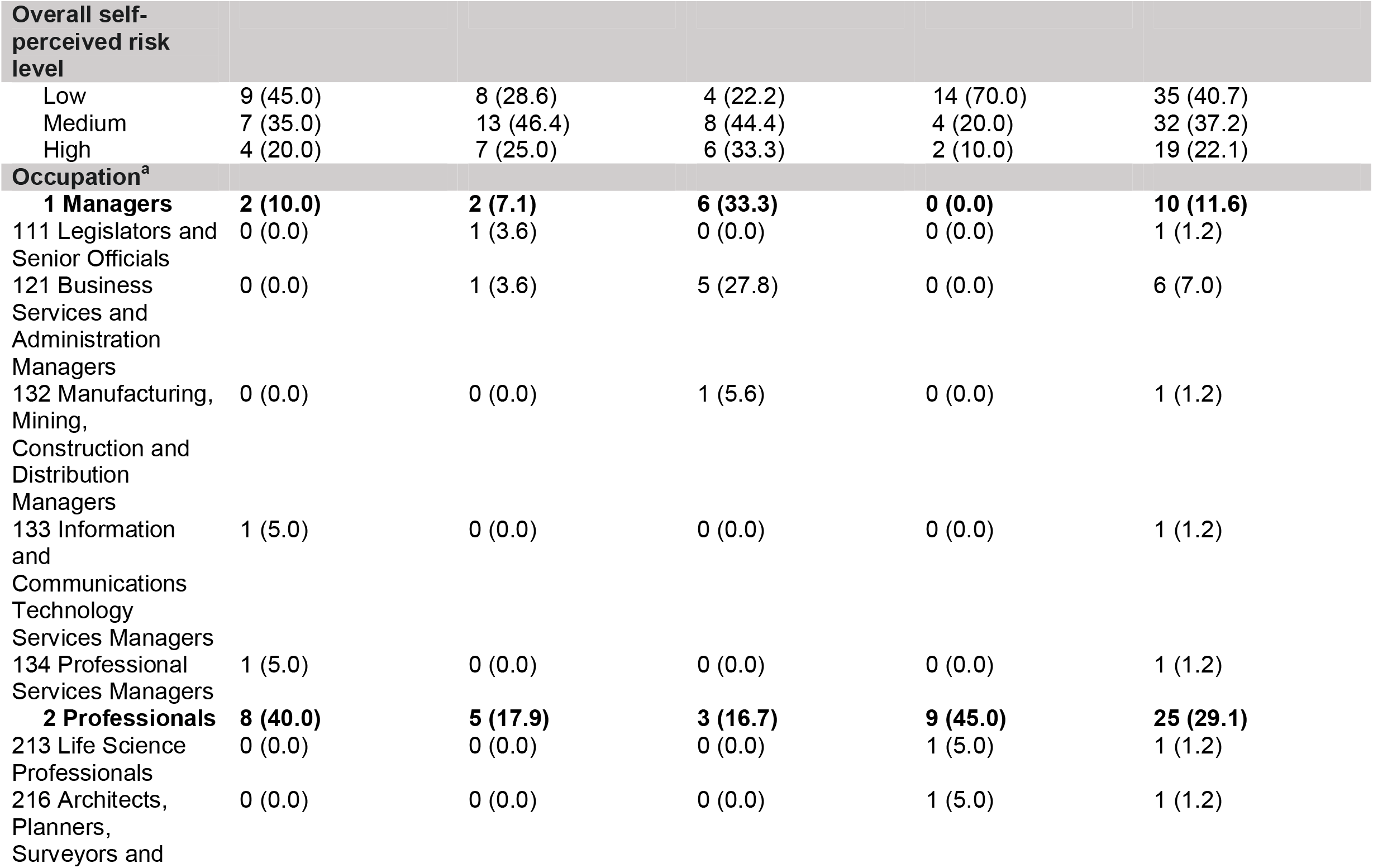
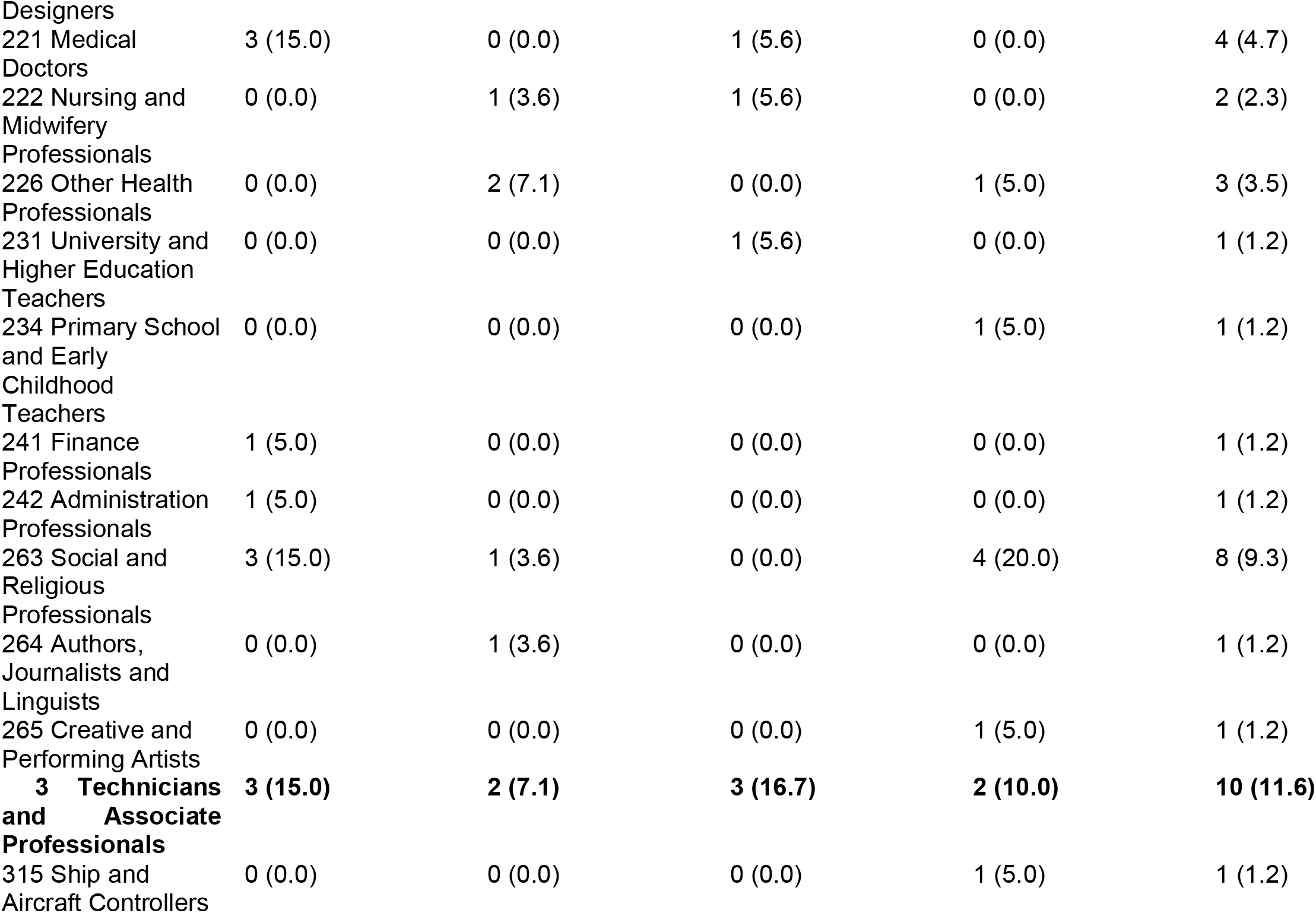
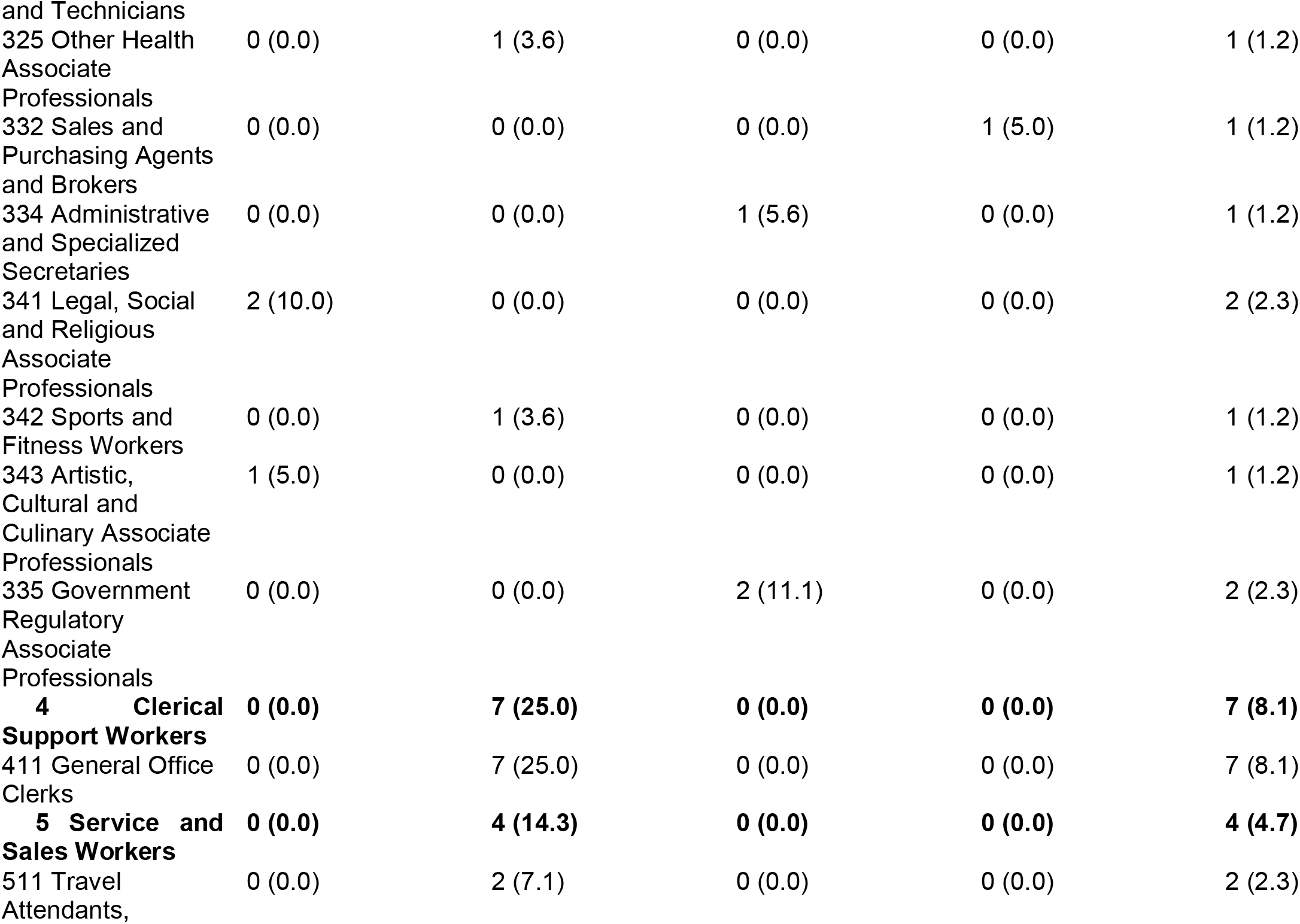
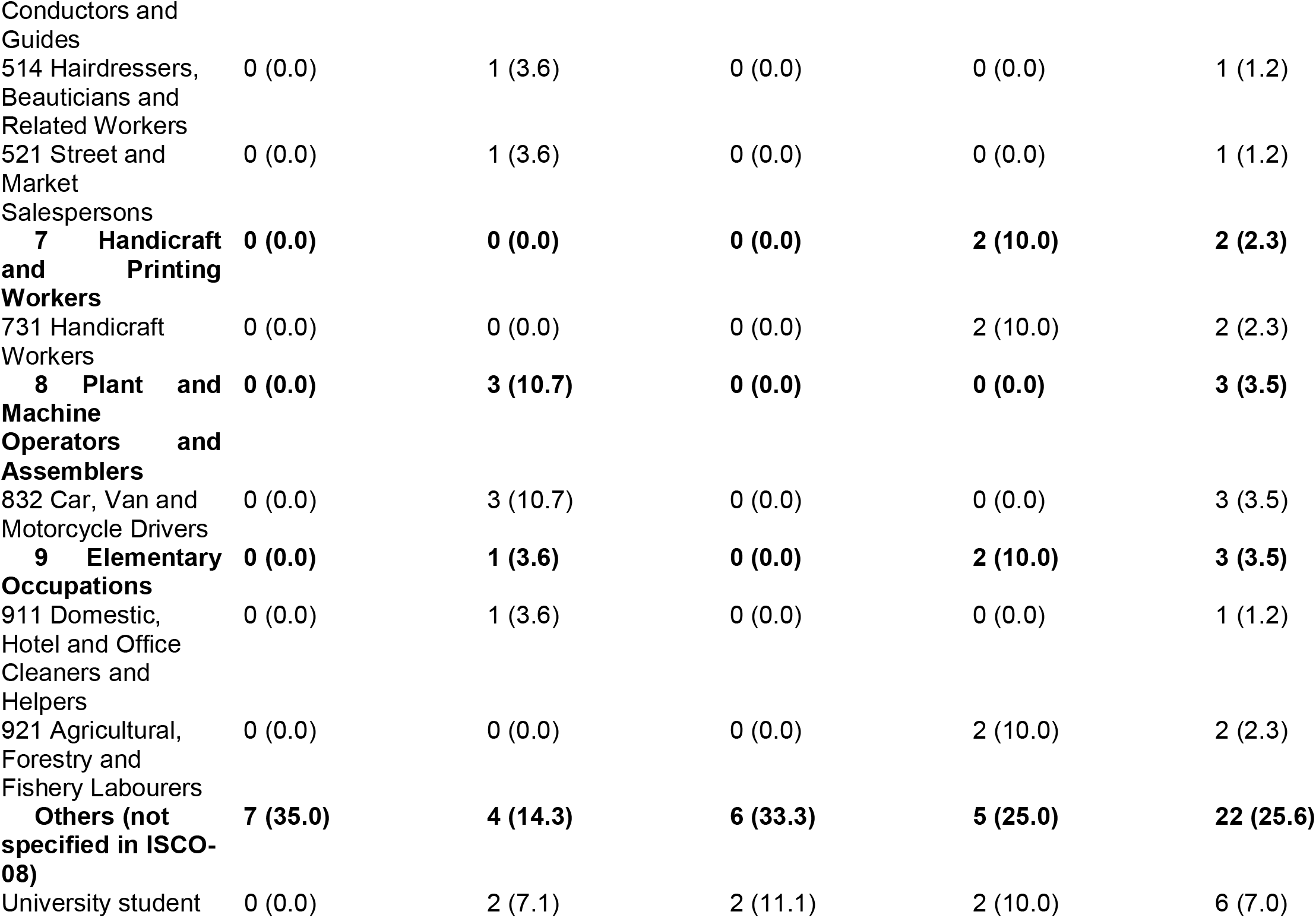
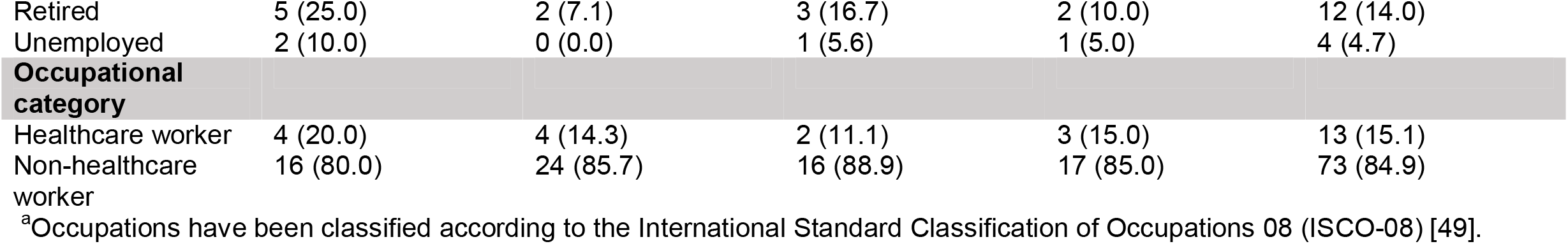
Participant characteristics.

### Data collection

The interview topic guide was developed collaboratively in a series of online meetings involving all interviewers from across the four countries. The topic guide was based on the research aims and protocol of the SEB-COV mixed-methods study [28]. Following pilot testing in each country, the guide was subsequently discussed and refined to focus on three broad areas, namely: (1) lived experiences and perceptions of COVID-19 measures (i.e., social isolation, social distancing, travel restrictions, quarantine); (2) wellbeing and mental health; (3) information, misinformation and rumours (for topic guide see [33]). In Thailand and Italy, topic guides were translated into Thai and Italian respectively, and interviews were conducted in these languages. In Malaysia and the UK, interviews were conducted in English.

Written informed consent was obtained electronically and permission was sought to audio record interviews. In Thailand and Malaysia, all interviews were recorded and transcribed. In Italy and UK, interviewers took detailed notes during the interview and wrote up summary scripts directly after, with salient quotes being transcribed verbatim from the recordings. The interview script method has been shown to produce levels of detail comparable to audio-recorded transcription [34, 35], and was chosen for reasons of time and resource efficiency. Because of data protection regulations preventing re-contacting of participants, transcripts and summaries were not returned to participants for validation and feedback. Interviews were scheduled at a time convenient to participants and lasted between 30-90 minutes.

As COVID-19 regulations prevented meeting face-to-face, the vast majority of interviews in all four countries were conducted by telephone or an online videoconferencing platform (Microsoft Teams, using video if possible), based on participants’ preferences. Following easing of local restrictions, one interview in Thailand and four interviews in Italy were conducted face-to-face at participants’ request, while adhering to local social distancing guidelines.

Only the researcher and participant were present during interviews and participants were encouraged to signal to the researcher if they felt that their privacy was compromised at any time (e.g., by the arrival of family members). No participants indicated this.

### Research team and reflexivity

In each country, interviews were conducted by a team of trained qualitative researchers (Thailand: BN, SR, TP; Malaysia: PKC, TS; Italy: GC, SS; UK: MLS, CMY; SR=male, all others=female), who began by introducing themselves and briefed participants about the aims of the study. Interviewers did not know participants before the study onset. All interviewers were residents in their study country, were affiliated with a national research institution, and fluently spoke the interview language.

### Data analysis

Data was collected iteratively, with emerging insights from each interview used to inform subsequent interviews, and continued until reaching ‘thematic saturation’, the point at which no salient new themes were deemed to emerge within each country [36]. Following completion of data collection, the Framework Method was used to analyse interview data, a tool commonly used in multi-disciplinary health research, offering “a systematic and flexible approach to analysing qualitative data”, particularly amongst large teams [30]. The Framework Method was chosen because of its efficiency and appropriateness for analysing data collected for pre-defined aims and objectives [37]. Based on the research aims and discussions within the cross-country research team, a broad, deductive set of thematic codes, including primary and secondary level codes, was developed.

The coding framework was tested, refined and subsequently applied to code data in each country. Coded data was then charted into the Framework matrix, which comprised three major themes (1. Views on public health measures; 2. Lived experience and impact of NPIs; 3. Coping strategies), and additional subthemes. Regular online meetings between the lead researcher (MLS) and country teams were used to review and discuss interpretations and coding of data, as well as charting of interview data into the Framework matrix. Finally, key findings for each theme were discussed and consolidated in online coding workshops between the lead researcher and country teams. These meetings supported a diversity of perspectives on data analysis, helping to strengthen reliability and validity of the findings [38].

### Ethical approval

Ethics approval was granted by Oxford Tropical Research Ethics Committee (OxTREC, reference no.520-20), covering all four countries. Additional ethics committee approval from Italy was not required for the study to be conducted there. In Thailand, additional approval was sought from the Faculty of Tropical Medicine Ethics Committee (FTMEC, ref: MUTM 2020-031-01). In Malaysia, additional approval was granted by the Medical Research and Ethics Committee (MREC), the Ministry of Health Malaysia (MOH ref: NMRR-20-595-54437 (IIR)), and the Universiti Tunku Abdul Rahman (UTAR) Scientific and Ethical Review Committee (SERC, ref: (U/SERC/63/2020).

## Results

Below we summarize key findings for each of the three major themes discussed. Supporting quotes which are referenced below (e.g., Q1, Q2, Q3 etc) are presented in Table 2.

**Table 2.**
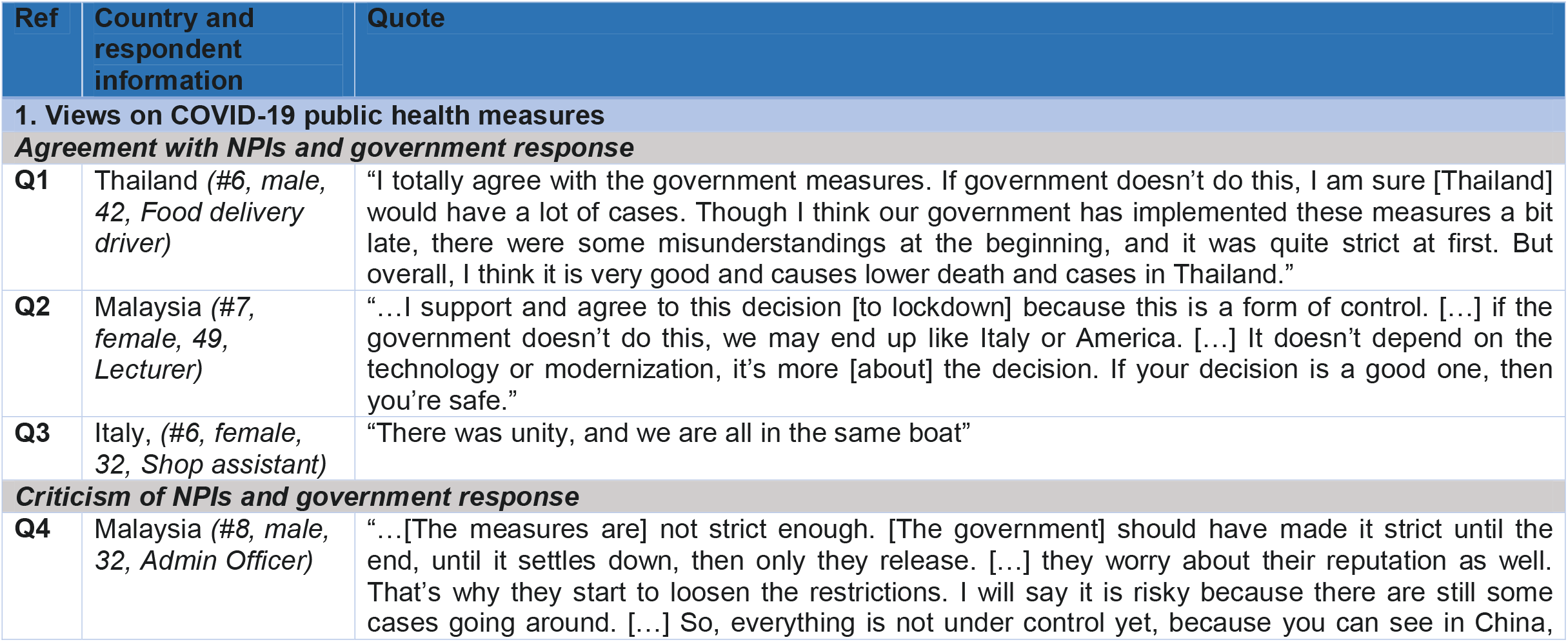
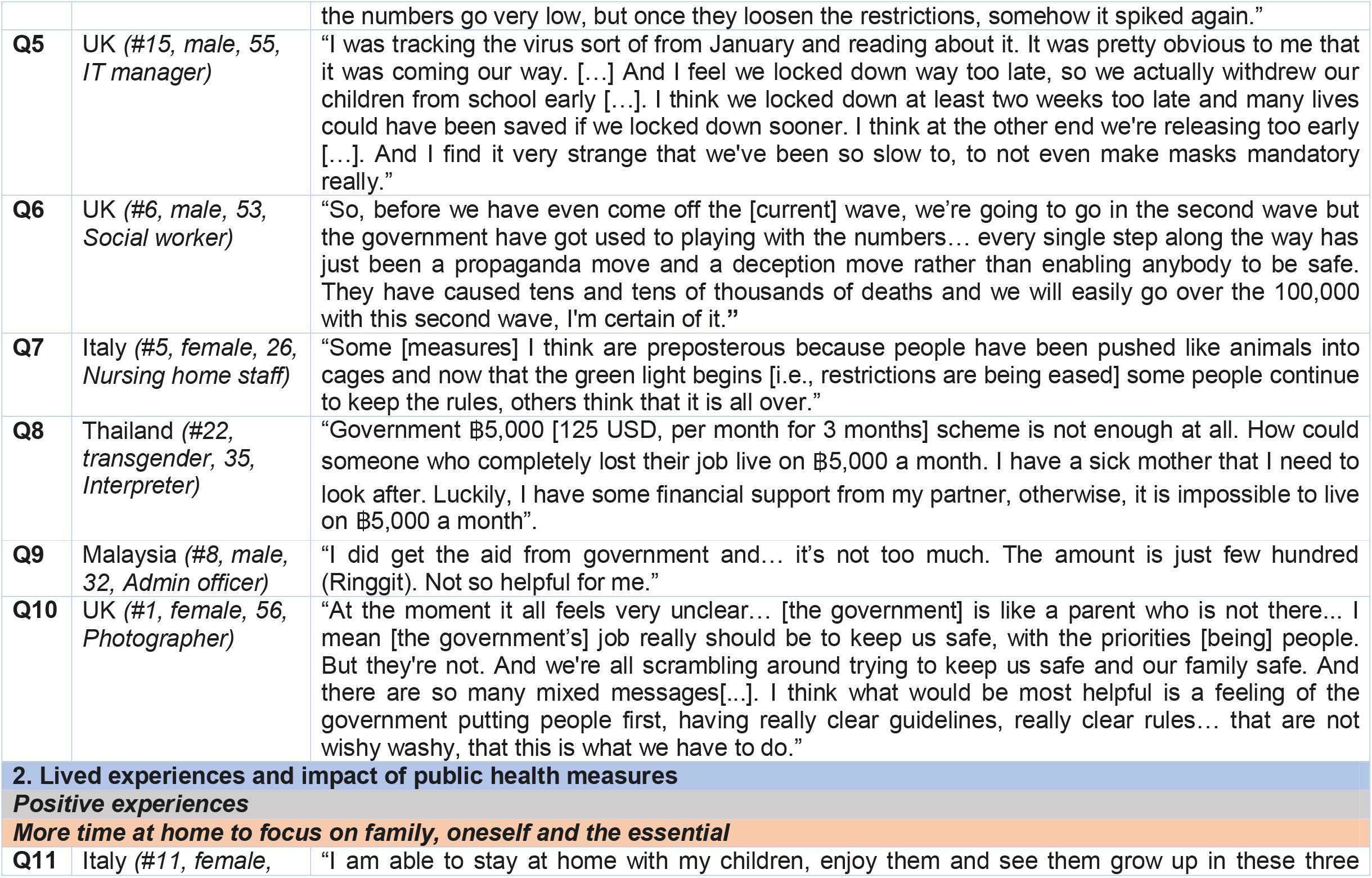
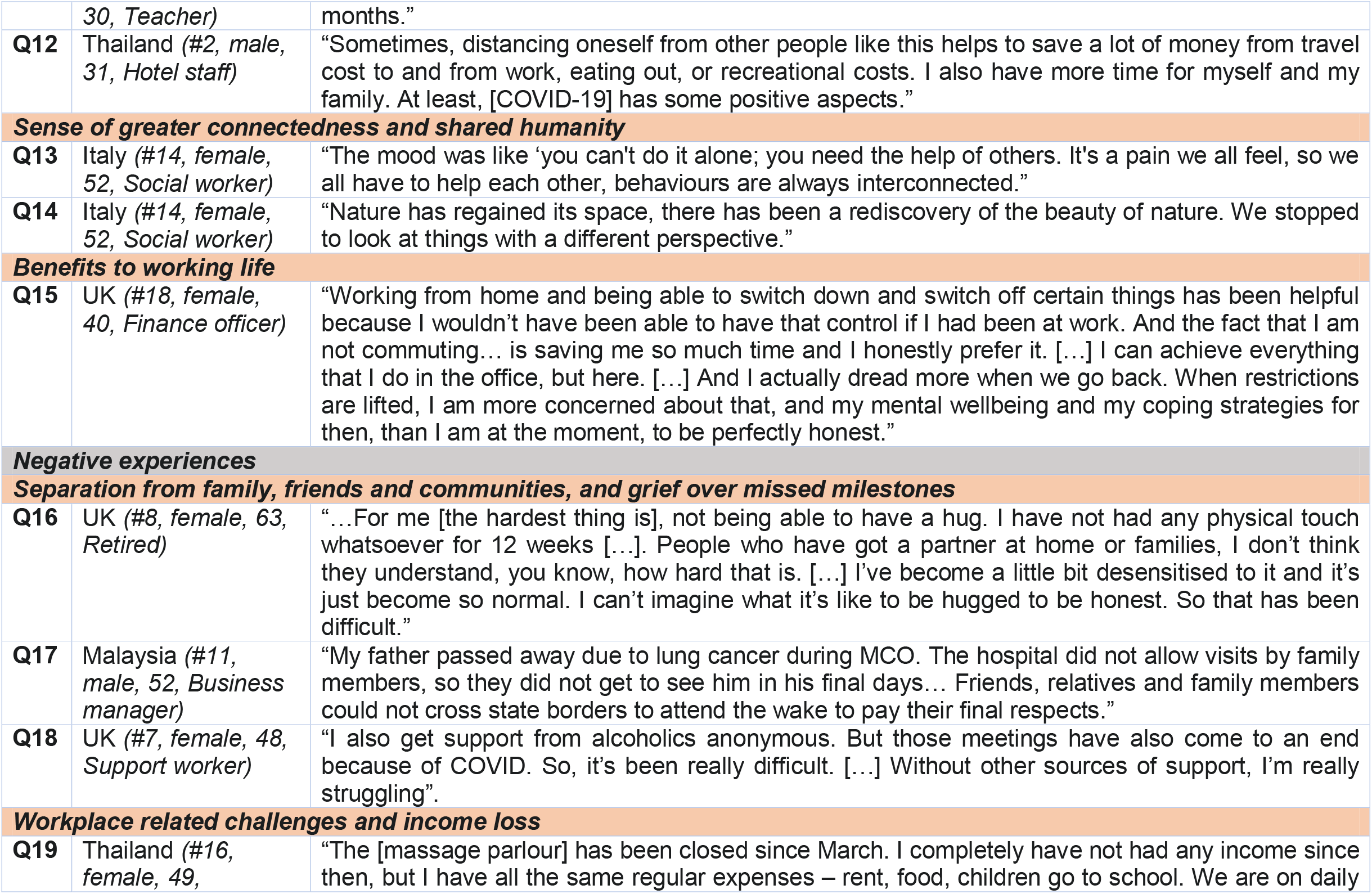
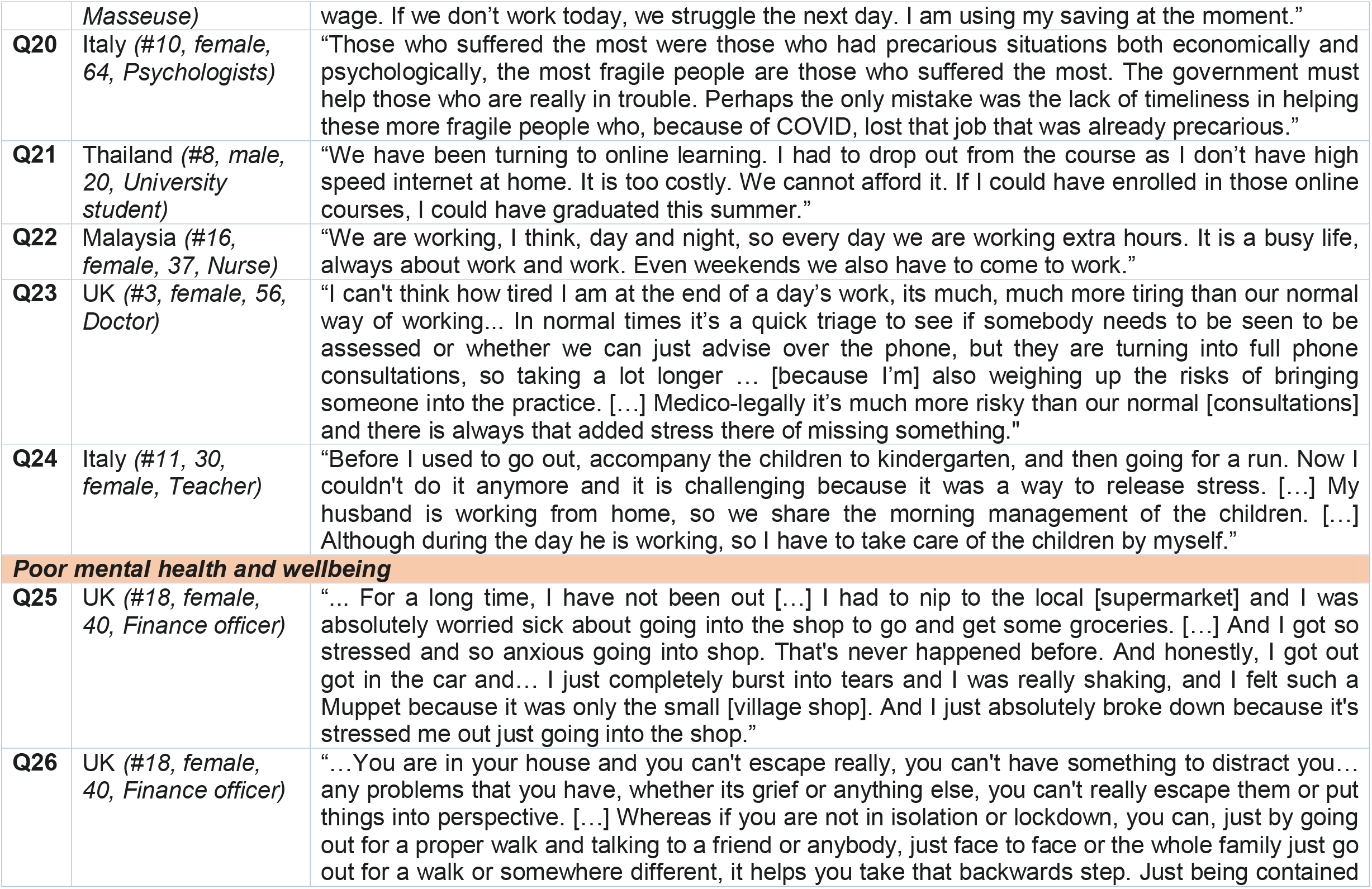
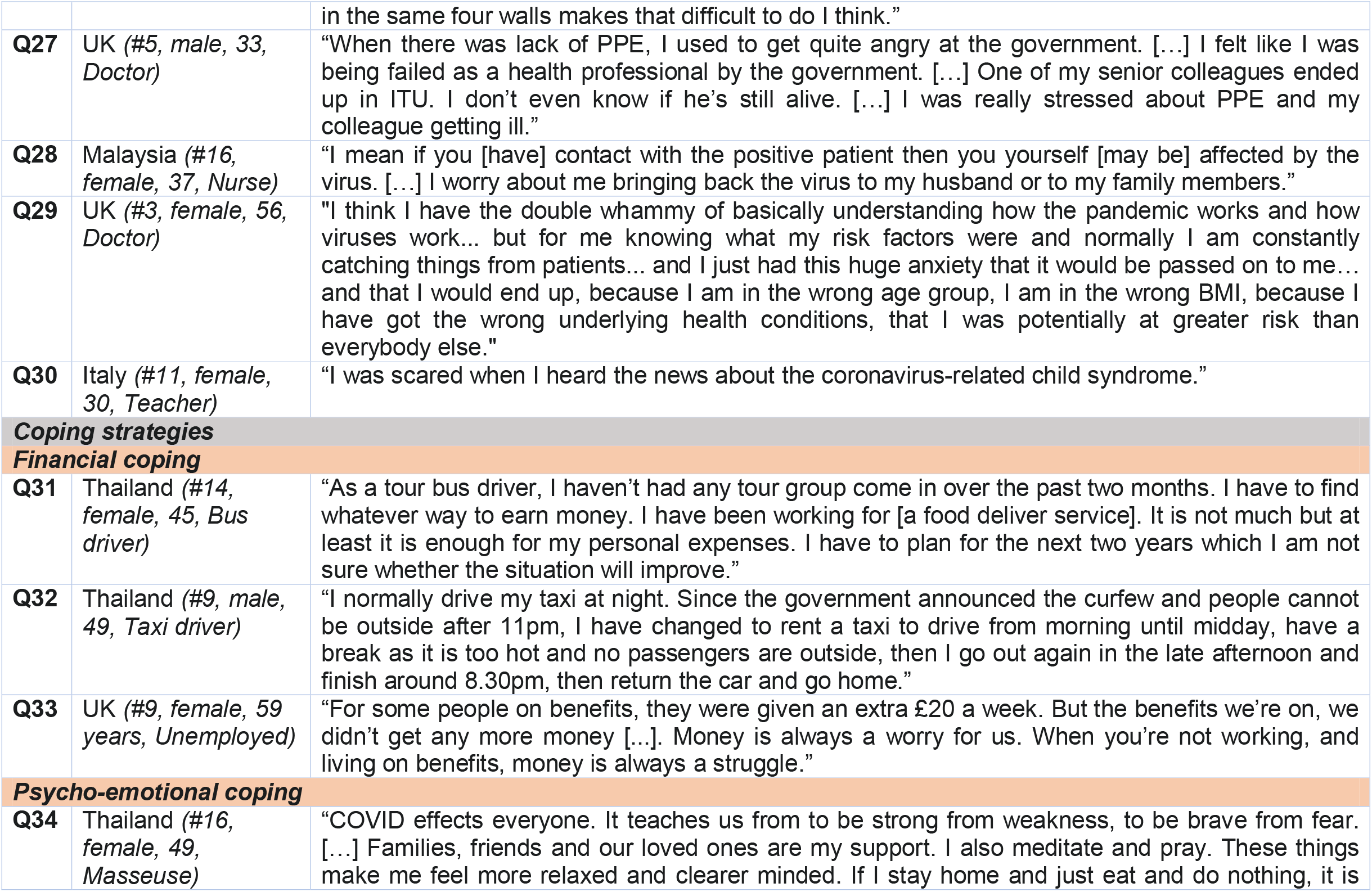
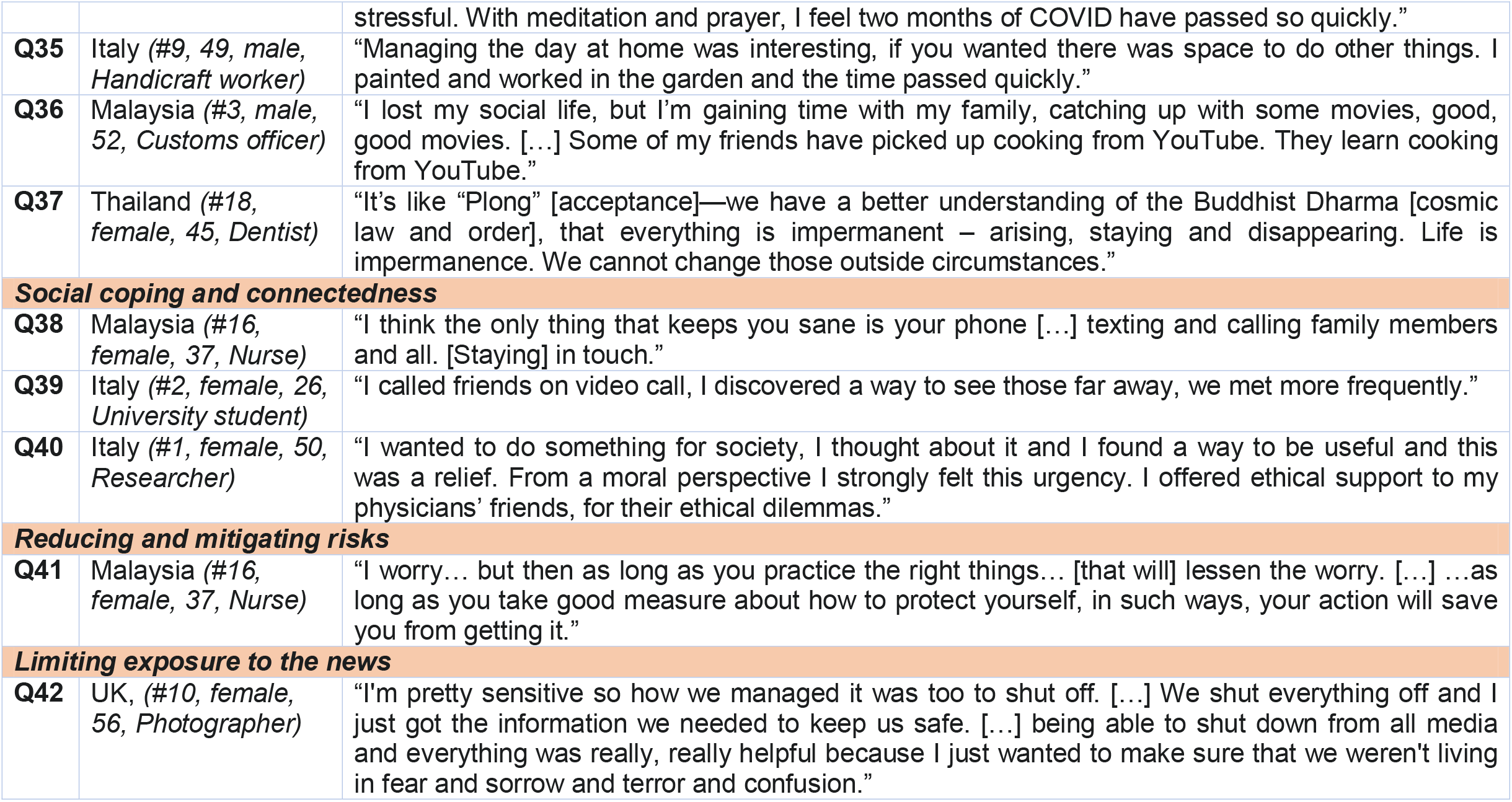
Supporting quotes from in-depth interviews conducted across four countries.

### Views on COVID-19 public health measures

Across the four countries, participants held complex and nuanced views about COVID-19 measures, which varied depending on participant’s personal circumstances, concerns and goals (e.g., staying safe/at home vs maintaining income/continuing to go out to work), and the extent to which NPIs were seen to facilitate or hinder these goals.

### Agreement with NPIs

Across countries, those who largely agreed with NPIs said that the measures, albeit being strict, were necessary to prevent community spread of COVID-19 (see Q1, Thailand). Many expressed feeling safer as a result of imposed NPIs. Some said their government had handled the pandemic well compared to other countries, thus deeming NPIs to be effective (see Q2, Malaysia). Such views were sometimes linked to narratives about the government doing their best amidst challenging circumstances and emphasised a sense of unity among the people, who found themselves “all in the same boat” (see Q3, Italy).

### Criticism of NPIs

Disagreement with government handling of COVID-19 ranged from mild criticisms about certain aspects of NPIs to some participants saying their government had failed to respond effectively. These participants believed that current NPIs were insufficient and “not strict enough” (see Q4, Malaysia) to contain further spread. For example, some said their government had done too little and “locked down way too late” (see Q5, UK), that rules lacked enforcement by the police and criticised premature easing of restrictions, raising fears about a “second wave” (see Q6, UK).

Contrarily, others felt that NPIs were too strict “because people have been pushed like animals into cages” (see Q7, Italy). Some Italian participants also believed that measures should have been implemented regionally, not nationally. In Thailand and Malaysia, some participants said that measures like curfews, travel restrictions and boarder closures threatened livelihoods, such as in tourism and for migrant workers, and that financial support from the government was insufficient (see Q8, Thailand; Q9, Malaysia).

Many participants also reported confusion and a lack of clarity in relation to NPI rules and regulations in their country. Particularly those in Italy and the UK commonly voiced frustration about the “mixed messages” and lack of clear NPIs guidelines from the government, leaving many to feel unsafe (see Q10, UK).

### Lived experiences and impact of public health measures

Most participants reported that NPIs impacted their lives in negatives ways. However, many also discussed inadvertent positive experiences, opportunities or ‘silver linings’, which had resulted from the COVID-19 measures. The extent of both positive and negative experiences varied considerably between participants, based on personal circumstances, particularly financial security, and across countries. Participants also discussed coping strategies adopted to mitigate the negative impacts of the measures. Fig 2 shows a summary of key findings.

**Fig 2.**
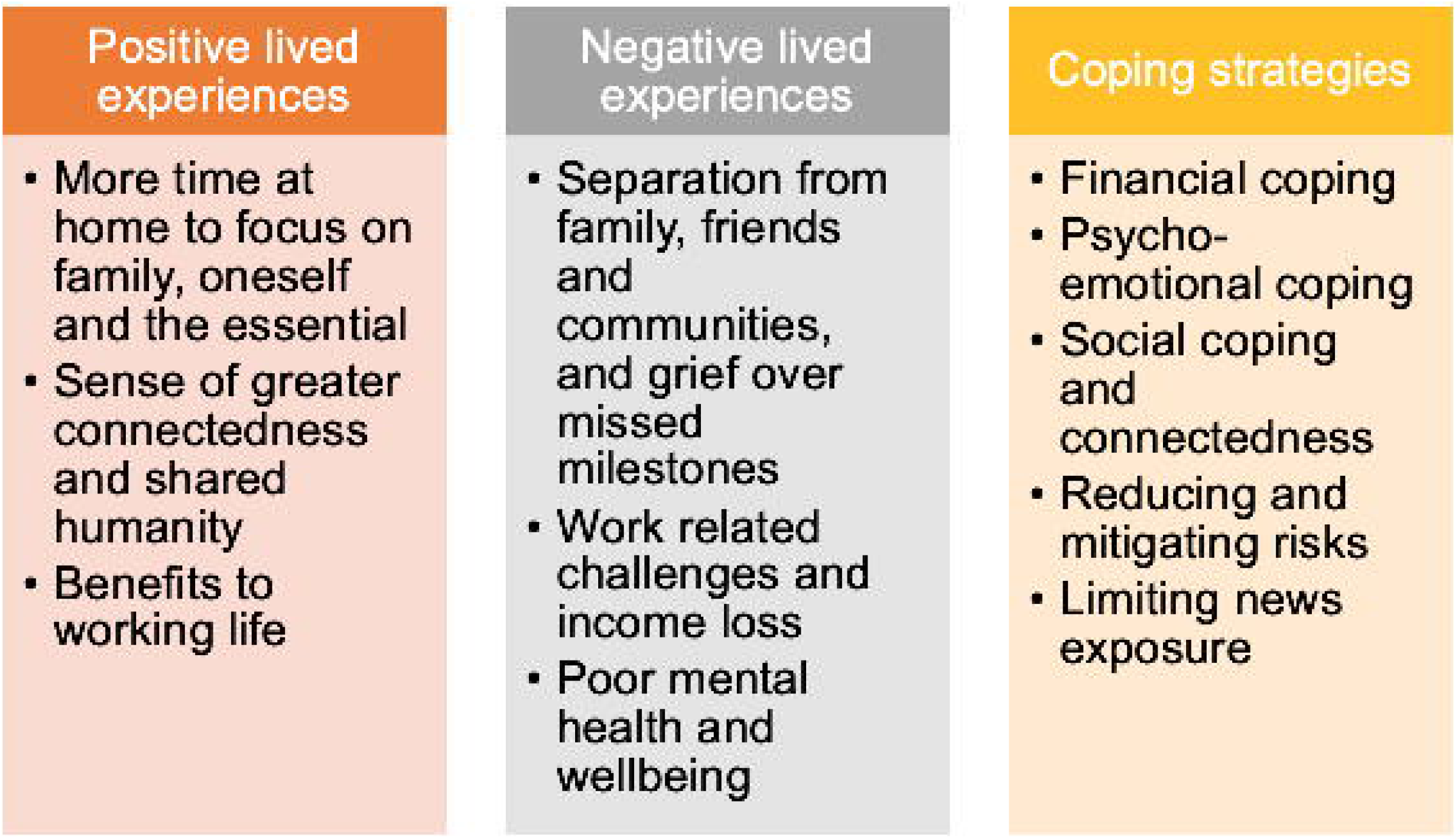
Lived experiences and impact of living under COVID-19 public health measures in four countries. Summary of key positive experiences, negative experiences and coping strategies.

### Positive experiences

#### More time at home to focus on family, oneself and the essential

Having more time to spend at home was a key positive experience reported across the four countries. Many compared the constant busyness of their ‘normal’ lives with the quiet and stillness during lockdown, prompting them to focus on what they felt was most essential in life. For some, this facilitated more time with family, extended parental leave, helping children with schoolwork, or reconnecting remotely with family and friends (see Q11, Italy). Others appreciated having more time to themselves, learn new skills, rest and reflect, engage in spiritual practice, or establish a healthier lifestyle, through exercise and cooking. Some enjoyed feeling less pressure to socialise and also said they “save[d] a lot of money” while staying at home (see Q12, Thailand).

#### Sense of greater connectedness and shared humanity

Across countries, some participants reported experiencing a greater sense of social support and solidarity within their communities during lockdown. Many said local support and volunteering initiatives that had sprung up in light of the community coming together, saying that the crisis had triggered a greater recognition of being “interconnected*”* and a sense of shared humanity (see Q13, Italy). Some participants in the UK and Italy additionally noted that the pandemic had made them feel more connected and appreciative of “the beauty of nature” (see Q14, Italy).

#### Benefits to working life

Some participants said that the measures had had a positive impact on their working lives. Across countries, some participants welcomed more flexible hours, home working, the lack of or reduced commute, with less traffic and pollution, while some also reported increased productivity and decreased work-related stress (see Q15, UK). A few deviant cases, participants also mentioned new business opportunities resulting from restrictions, such as setting up food delivery businesses at home.

### Negative experiences

#### Separation from family, friends and communities, and grief over missed milestones

Participants across all countries described their separation from friends and family as the most challenging aspect of NPIs. Many felt isolated and lonely due to prolonged physical separation from their loved ones, especially participants who lived alone (see Q16, UK).

Separation and isolation were described as most challenging when participants were unable to communally celebrate significant milestones, such as birthday, weddings, religious ceremonies, holidays or funerals (see Q17, Malaysia). Across countries, many described a feeling of ‘missing out’ on important life events, with this being particularly poignant among younger and older participants, for instance around experiencing grandchildren’s development or being involved in activism.

The closure of recreational facilities, such as cinemas and gyms, or social support services and community organisations, such as mental health support groups or religious services, was perceived as particularly challenging by some (see Q18, UK).

#### Work related challenges and income loss

Many participants across countries described the negative impacts of NPIs on their work. Thai and Malaysian participants, in particular, and to a lesser extent UK and Italian participants, said that the fear of, or actual loss of their job or income was a grave consequence of the restrictive measures (see Q19, Thailand).

Some participants also worried about the impact of NPIs on society and the economy more broadly, expressing particular concerns about mass unemployment and negative consequences for those in “precarious situations, both economically and psychologically” (see Q20, Italy).

Many who continued working reported challenges due to their altered work situation, including challenges related to poor connectivity (see Q21, Thailand), difficulties concentrating while home-working, increased workloads, and heightened work-related stress and anxiety, particularly among HCW (see Q22, Malaysia; Q23, UK).

In Malaysia and Thailand, participants complained that travel restrictions and curfews caused pressure on their work schedules and livelihoods. Some young professionals expressed concerns about disruptions to their career progression. Finally, women in particular described the increased burdens of unpaid care and domestic work during lockdown (see Q24, Italy).

#### Poor mental health and wellbeing

The negative impact of lockdown measures on mental health and wellbeing was a consistent theme across countries. Several participants reported low mood, grief, fatigue, restlessness, boredom, guilt, feeling overwhelmed and a lack of control as a result of the changes to their personal, family and work lives. For many participants, concerns about contracting COVID-19 resulted in strong fear and anxiety, including around leaving the house or doing shopping (see Q25, UK). At the same time, some described how being stuck at home made it difficult to “escape” these negative thoughts and feelings or “put things into perspective” (see Q26, UK).

Reports of COVID-related stress and anxiety were heightened among those who considered themselves or a family member to be vulnerable, and among HCW. Frontline HCW frequently complained about a lack of personal protective equipment (PPE) or changing PPE guidelines, causing considerable fears about their own and their household member’s health (see Q27, UK; Q28, Malaysia), particularly among those with underlying health conditions (see Q29, UK).

Future uncertainties resulting from NPIs, such as when and how the measures would be eased and the impact on job prospects, was reported as a major stressor on mental health. Parents also commonly worried about the long-term impact of the measures on their children (see Q30, Italy).

### Coping strategies

Across countries, participants reported various ways of coping with the impacts of the COVID-19 measures.

### Financial coping

Many participants described their efforts to cope with financial challenges and income loss resulting from COVID-19 restrictions. For example, some participants tried to reduce spending where possible, while others took on additional work, such as starting a home business or taking on additional work (see Q31, Thailand).

In Thailand and Malaysia, informal sector workers described adapting working schedules around imposed curfews and travel restrictions (see Q32, Thailand). Participants in the UK, Italy and Thailand, who received financial support from their governments described this as helpful but insufficient to make up for lost income (see Q33, UK).

### Psycho-emotional coping

In adapting to the ‘new normal’, many participants actively tried to maintain wellbeing amidst difficult life circumstances, with similar coping strategies being reported across countries. These included engaging in religious and spiritual practices (e.g., meditation, prayer) (see Q34 Thailand); establishing daily routines around self-care, work and childcare (e.g., regular exercise, sleep, getting dressed as if going to work, home schooling schedules); maintaining relationships, social connections and recreational activities from home (e.g., using online media) (see Q35, Malaysia); and making home improvements (e.g., tidying, doing home repairs, gardening) (see Q36, Italy).

As a way of coping, some Thai participants, in line with Buddhist principles spoke about intentionally trying to cultivate attitudes of ‘acceptance’ and ‘letting be’ (i.e., “*Plong*”), rather than focusing on the uncertainty surrounding pandemic restrictions (see Q37, Thailand).

### Social coping and connectedness

Across countries, staying connected with friends, family and peers through shifting social routines online and via telephone was considered essential for keeping “sane” during prolonged physical separation from loved ones and for maintaining a sense of connectedness (see Q38, Malaysia; Q39 Italy).

Some participants felt that a renewed sense of solidarity and communities coming together helped to counter feelings of loneliness and isolation. Several reported “want[ing] to do something for society” (see Q40, Italy), saying that volunteering initiatives or providing personal support to family, friends, neighbours and colleagues helped them to experience a sense of purpose during this difficult time.

### Reducing and mitigating risks

Across countries, many participants reported taking steps to reduce potential exposure to COVID-19, including staying at home; switching to online grocery shopping or designating one person per household to do in-person shopping; using PPE like hand sanitiser, gloves, face masks and shields; and avoiding close contact with others when in public. Particularly HCW reported that having access to PPE reduced fears and increased their sense of control (see Q41, Malaysia).

### Limiting exposure to the news

Across countries, many participants reported feeling distressed by the constant exposure to COVID-19 news, which some felt were unclear, untrustworthy and politicised. As such, in an effort to balance their desire to stay informed and to manage the anxiety evoked by the news, many participants said they deliberately limited their daily news exposure (see Q42, UK).

## Discussion

This study provides insight into the lived experiences of NPIs among the public, including HCW across four countries during the first wave of the COVID-19 pandemic, an unprecedented time in history. Despite considerable variation of individual circumstances, there was surprising similarity of experiences across countries, including around personal, social, familial, professional, financial, religious and spiritual life domains. This study unusually highlights the positive silver linings and coping strategies that individuals adopted, alongside the better understood negative psychosocial and economic impacts.

Across countries, participants generally had strong views, either supportive or critical, of their government’s handling of the pandemic, with few expressing neutral views. Criticisms of NPIs emphasised that measures were either not strict enough or too strict, thus threatening livelihoods. Overall, many expressed confusion and frustration at rapidly changing rules, echoing other research linking negative views of NPIs to mistrust of politics and negative perceptions of government transparency [20, 39]. Those supportive of NPIs emphasised the necessity of measures to prevent COVID-19 transmission, appealing to values of unity and solidarity. Across countries, community support and solidarity were also highlighted as helpful for coping with the psychosocial impacts of prolonged physical distancing and isolation. Public health messaging that emphasises shared values of solidarity, altruism and social responsibility are thus likely to enhance acceptability of NPIs, as has been shown for promoting the uptake of face coverings across several countries [40].

Most participants felt predominantly negatively affected by NPIs, including through income loss, work related challenges, separation from loved ones, grief over missed milestones, and widespread experiences of poor mental health. This supports existing qualitative evidence from the UK highlighting the negative impacts of social distancing and isolation, including economic, social, psychological, and emotional losses, such as loss of motivation, meaning and self-worth [20]. Our findings further support others showing that negative experiences were often amplified among HCW who faced greater work-related stress and exposure to COVID-19 risks [22, 23, 25, 41]. Furthermore, we highlight that experiences among HCW differed depending on their own levels of self-perceived COVID-19 risk, which resonates with research documenting significantly higher levels of COVID-19 infection among patient-facing HCW, compared to non-patient facing HCW [42].

Our findings additionally highlight inadvertent positive experiences and coping strategies emerging from this crisis, largely overlooked to date. Positive experiences included participants having more time to focus on their family, themselves and what they most valued; making positive changes to working routines; and feeling a greater sense of community. Many participants across countries rapidly adapted their lives by developing financial, psycho-emotional and social ways of adjusting to the ‘new normal’, and by reducing COVID-19-realted risks and exposure to anxiety provoking news. However, socio-cultural and economic factors importantly underpinned the extent to which participants were able to employ positive coping strategies. During this pandemic, some groups, including younger people, sexual and gender minorities and those with financial instability have been evidenced to use less productive coping strategies, like substance use and behaviour disengagement [43]. We support others who argue that, in order to ensure that COVID-19 interventions are equitable, the definition of who is considered vulnerable and ‘most-at-risk’ should be expanded to include social and cultural—not just biological—factors [44-46]. Further research on the lived experiences of coping with COVID-19 restrictions is therefore important for better understanding the socio-cultural factors that underpin differential susceptibility to experiencing adverse impacts of NPIs.

Importantly, understanding positive experiences and coping—and thereby shedding light on the full spectrum of lived experience—can illuminate and facilitate the public’s resilience in light of ongoing restrictions. This study suggests that efforts to maintain mental health services, community and religious support groups remotely; ensuring availability of PPE; and offering financial compensation for lost income are critical ingredients to support better coping across countries. Furthermore, identifying socially vulnerable groups in each context and accounting for particular socio-cultural needs, through community engagement and social science research, will be essential for policy makers to promote positive coping strategies and reduce the negative and unequal impacts of NPIs [19].

This study is limited in three main ways. Firstly, although we aimed to sample a diverse group, there was less variety in socio-economic status than desired. This was due to rapid recruitment, and interviews being conducted online or by phone, restricting access of those from lower socio-economic groups with less access to these technologies. Additionally, due to data protection regulations, only Microsoft Teams was approved for online data collection in this study, ruling out more accessible and widespread platforms like WhatsApp or Facebook Messenger, likely to have facilitated greater diversity among participants. Despite this, through purposive sampling we managed to include participants from a range of age groups, occupations, educational backgrounds and self-perceived risk levels, including HCW. Secondly, social desirability bias [47] may have led participants to represent themselves as overly adherent to and supportive of NPIs, and avoided revealing more extreme views (e.g., beliefs in conspiracy theories). Thirdly, while this study compared lived experiences of NPIs across countries, future research would benefit from investigating intra-country differences, including comparisons between urban and rural residents [48].

A strength of this study is that it is the first cross-country qualitative study to provide evidence on the impact of NPIs on lived experience of the public, including HCW, which can help to inform the COVID-19 policy response. Additionally, the use of remote interviews allowed for nationwide recruitment across the four countries, circumventing barriers of geographical distance, while also potentially offering a greater sense of safety and anonymity, which may have increased participation from individuals who may be reluctant to participate in-person.

## Conclusion

Findings from this study underscore the need for COVID-19 policies and interventions to pay greater attention to the differential impact of NPIs on different population groups. Supporting individuals’ coping is an important strategy for mitigating the adverse impact of NPIs and interventions should focus on socially vulnerable groups who experience greater negative impacts and are less able to cope. These are likely to include individuals facing socio-economic deprivation and other marginalised populations, such as sexual and gender minorities and migrant workers. We emphasise that across countries, declines in mental health as a result of NPIs appear widespread and common. Efforts to counter this trend are thus critical: policy makers must prioritise populations physical health—i.e., protection from COVID-19 infection—while not losing sight of their mental health.

## Supporting information

disclosure statement

COREQ checklist

## Data Availability

All relevant data are within the manuscript and its Supporting Information files. Complete interview data cannot be made publicly available due to ethical and legal reasons. Upon reasonable request, a list of condensed meaning units or codes can be made available on request to the MORU Data Access Committee (see link https://www.tropmedres.ac/units/moru-bangkok/bioethics-engagement/data-sharing).

## Acknowledgements

We would like to thank all participants across the four countries who agreed to take part in this research.

